# Multiethnic Validation of Artificial Intelligence-Enhanced Electrocardiographic Image Analysis in Detecting Cardiac Structural and Functional Abnormalities: A UK Biobank Study

**DOI:** 10.1101/2025.07.08.25331159

**Authors:** Yerin Kim, Haemin Lee, Hong-Mi Choi, In-Chang Hwang, Joonghee Kim, Ji Hyun Lee, Il-Young Oh, Heesun Lee, Su-Yeon Choi, Tae-Min Rhee, Youngjin Cho

## Abstract

**Background:** Although artificial intelligence–enhanced electrocardiography (AI-ECG) has shown promise in detecting cardiac abnormalities, large-scale validation against cardiac magnetic resonance (CMR) parameters in a multiethnic general population remains limited.

**Methods:** In 38,804 UK Biobank participants with paired ECG and CMR data, we evaluated six AI-ECG models: four targeting functional abnormalities (left and right ventricular dysfunction [QCG-LVD and QCG-RVD], global longitudinal strain [ECG-LVGLS and ECG-RVGLS]) and two targeting structural abnormalities (left ventricular hypertrophy [AI-ECG-LVH] and left atrial enlargement [AI-ECG-LAE]). Abnormalities were defined as the top 1% extreme values of each CMR-derived parameter.

**Results:** The AI-ECG models demonstrated robust diagnostic performance. For left ventricular dysfunction, the AUCs were 0.887 (QCG-LVD) and 0.896 (ECG-LVGLS); for right ventricular dysfunction, the AUCs were 0.778 (QCG-RVD) and 0.825 (ECG-RVGLS). In a sub-cohort with available corresponding CMR data (n=21,267), AUCs were 0.824 for AI-ECG-LVH and 0.883 for AI-ECG-LAE. Subgroup analyses showed consistent performance across demographic and clinical groups, with enhanced accuracy observed in older individuals, males, and those with hypertension.

**Conclusion:** In this large-scale, population-based study, AI-ECG demonstrated strong performance in detecting structural and functional cardiac abnormalities defined by CMR. These findings support the potential utility of AI-ECG as a scalable screening tool for cardiovascular disease in general populations.

## Introduction

Electrocardiography (ECG) is a widely accessible, non-invasive, and cost-effective tool for evaluating cardiovascular health. Recent advances in artificial intelligence (AI) have significantly expanded its clinical utility.^1–3^ AI-enhanced ECG (AI-ECG) enables the prediction of structural and functional cardiac abnormalities that have typically required advanced imaging modalities such as echocardiography or cardiac magnetic resonance imaging (CMR). By leveraging the ubiquity of ECG, AI-ECG offers a scalable strategy for early detection and screening of cardiovascular disease, potentially reducing dependence on resource-intensive imaging tools.

Our group has previously developed and validated deep learning based AI-ECG models that analyze ECG images to generate digital biomarkers for various cardiac conditions.^4–7^ These models rely on image-based ECG input rather than raw signal data, offering flexibility in clinical implementation and demonstrating high diagnostic performance. However, despite their promising performance, further validation is necessary to assess their generalizability in broader populations, particularly across diverse ethnicities and outside of hospital-based cohorts.

This study aims to validate previously developed AI-ECG models in a general population from the UK Biobank, comprising over 38,000 participants with paired ECG and CMR data.

## RESULTS

### Baseline Characteristics

Baseline characteristics of the study population are summarized in Table 1. The mean age of participants was 64.2 ± 7.8 years, with 48.2% being male. Comorbidities included hypertension in 32.9%, diabetes mellitus in 5.9%, and dyslipidemia in 24.1%. The mean left ventricular ejection fraction (LVEF) measured by CMR was 59.5 ± 6.1%, and the mean right ventricular ejection fraction (RVEF) was 57.3 ± 6.2%.

**Table 1.**
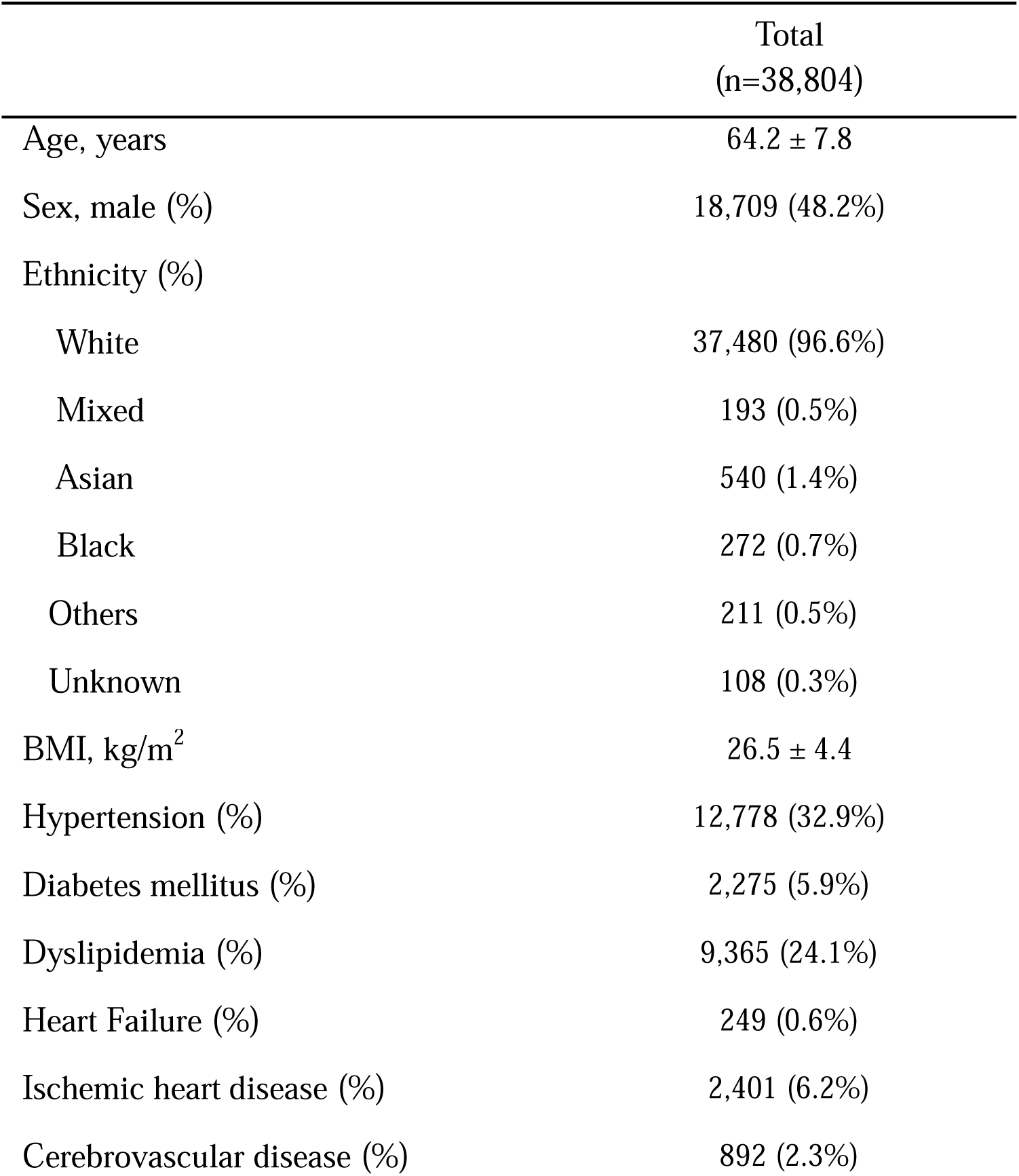
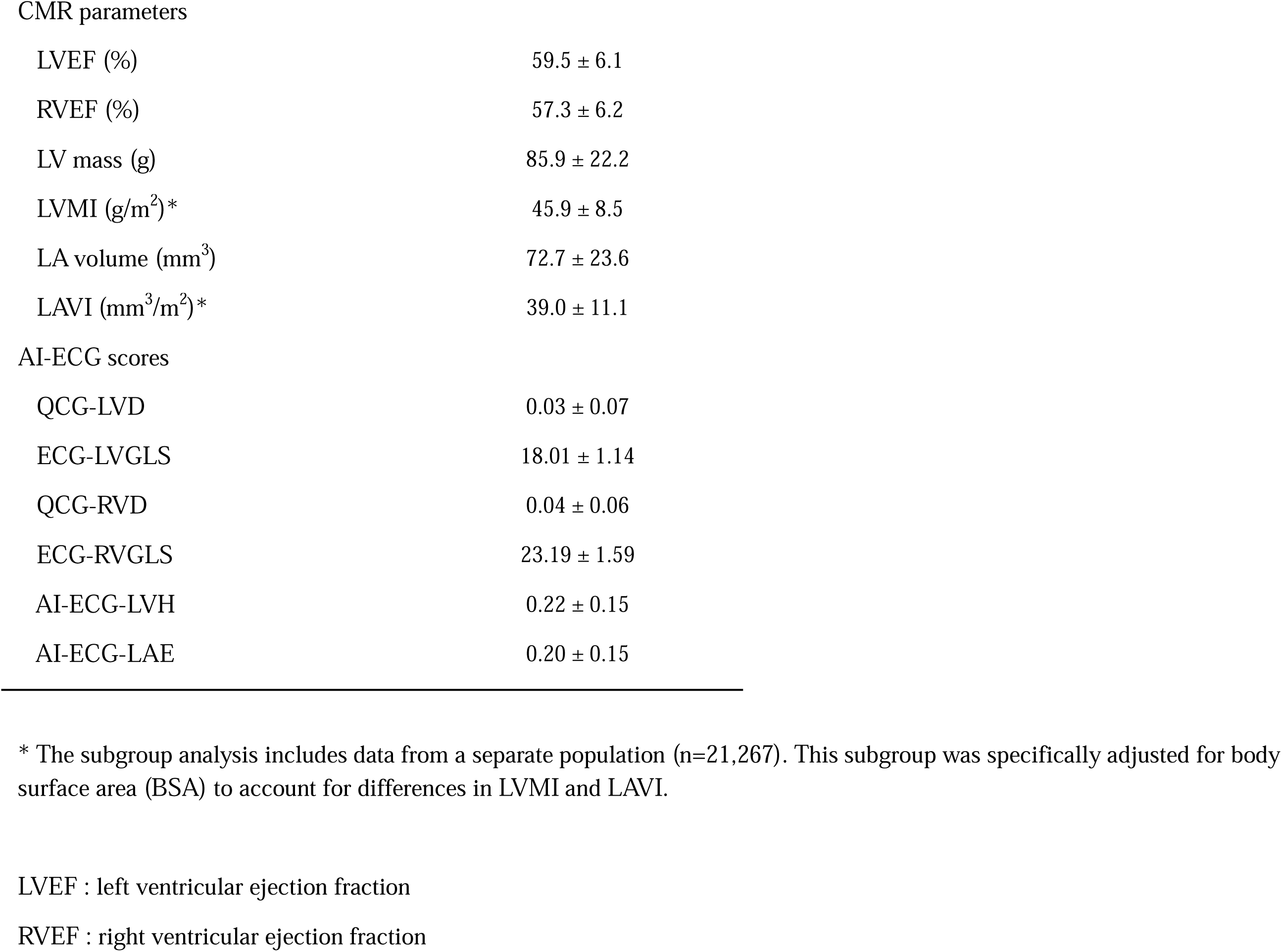
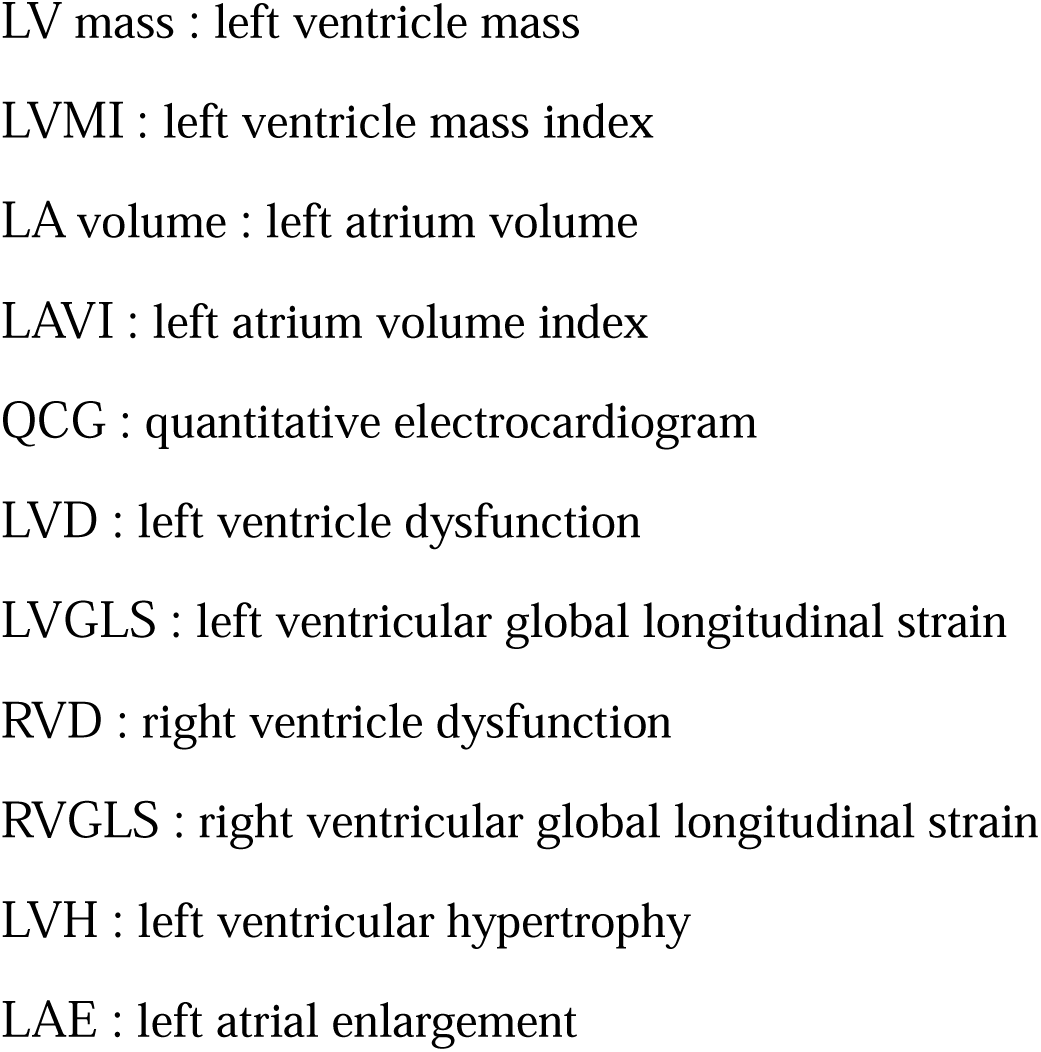
Baseline characteristics of the overall study population.

### AI-ECG Performance for Detecting Left and Right Ventricular Dysfunction

Table 2 presents baseline characteristics stratified by the presence of left ventricular dysfunction (LVD) and right ventricular dysfunction (RVD). Participants with LVD or RVD were generally older and had a higher prevalence of comorbid conditions compared to those without dysfunction. In particular, hypertension was more prevalent in both LVD and RVD groups. Individuals with LVD had higher rates of diabetes mellitus and ischemic heart disease, while those with RVD more frequently had heart failure.

**Table 2.**
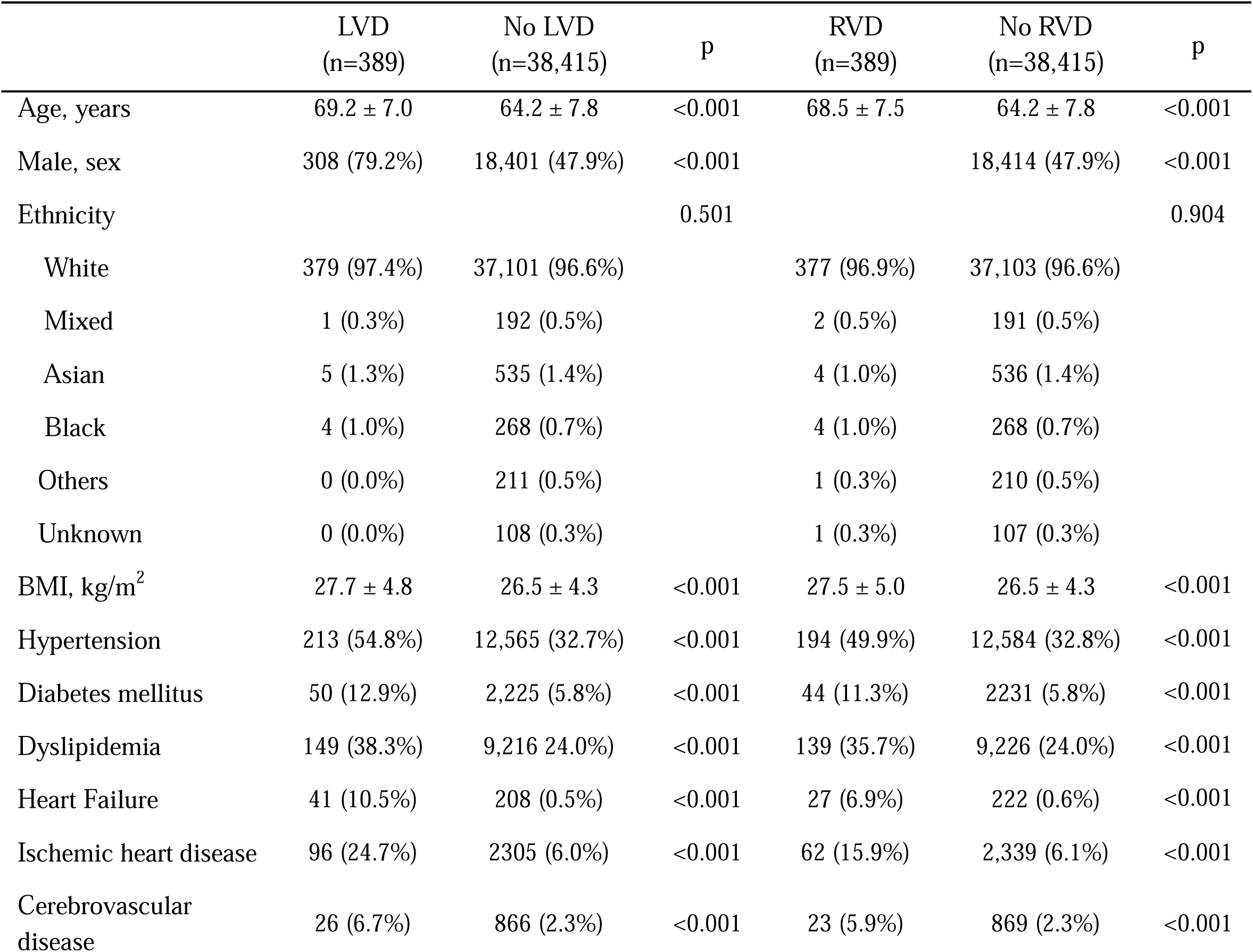
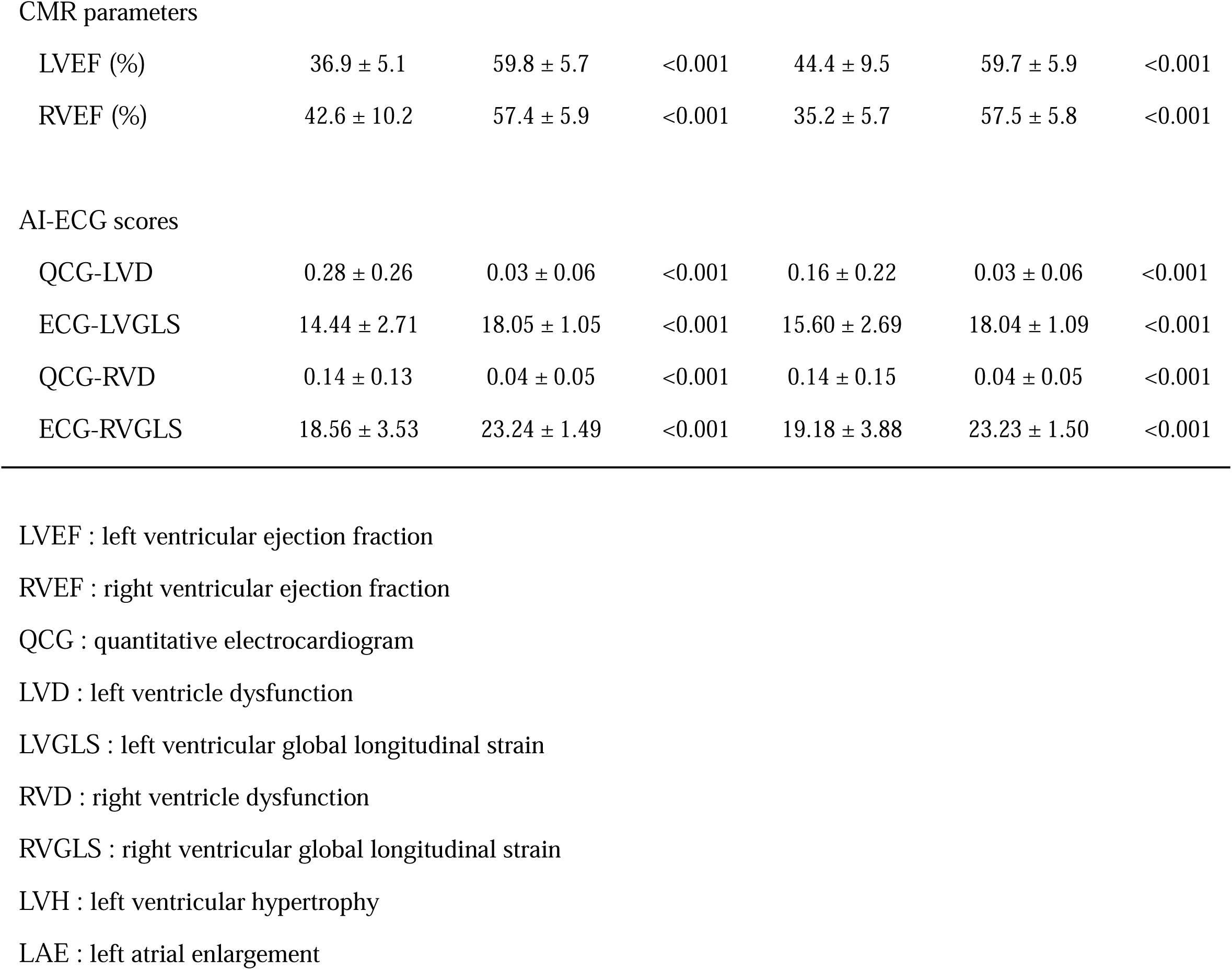
Baseline characteristics according to the presence of left and right ventricular dysfunction.

|  | LVD<br>(n=389) | No LVD<br>(n=38,415) | p | RVD<br>(n=389) | No RVD<br>(n=38,415) | p |
| --- | --- | --- | --- | --- | --- | --- |
| Age, years | 69.2 ± 7.0 | 64.2 ± 7.8 | <0.001 | 68.5 ± 7.5 | 64.2 ± 7.8 | <0.001 |
| Male, sex | 308 (79.2%) | 18,401 (47.9%) | <0.001 |  | 18,414 (47.9%) | <0.001 |
| Ethnicity |  |  | 0.501 |  |  | 0.904 |
| White | 379 (97.4%) | 37,101 (96.6%) |  | 377 (96.9%) | 37,103 (96.6%) |  |
| Mixed | 1 (0.3%) | 192 (0.5%) |  | 2 (0.5%) | 191 (0.5%) |  |
| Asian | 5 (1.3%) | 535 (1.4%) |  | 4 (1.0%) | 536 (1.4%) |  |
| Black | 4 (1.0%) | 268 (0.7%) |  | 4 (1.0%) | 268 (0.7%) |  |
| Others | 0 (0.0%) | 211 (0.5%) |  | 1 (0.3%) | 210 (0.5%) |  |
| Unknown | 0 (0.0%) | 108 (0.3%) |  | 1 (0.3%) | 107 (0.3%) |  |
| BMI, kg/m<sup>2</sup> | 27.7 ± 4.8 | 26.5 ± 4.3 | <0.001 | 27.5 ± 5.0 | 26.5 ± 4.3 | <0.001 |
| Hypertension | 213 (54.8%) | 12,565 (32.7%) | <0.001 | 194 (49.9%) | 12,584 (32.8%) | <0.001 |
| Diabetes mellitus | 50 (12.9%) | 2,225 (5.8%) | <0.001 | 44 (11.3%) | 2,231 (5.8%) | <0.001 |
| Dyslipidemia | 149 (38.3%) | 9,216 (24.0%) | <0.001 | 139 (35.7%) | 9,226 (24.0%) | <0.001 |
| Heart Failure | 41 (10.5%) | 208 (0.5%) | <0.001 | 27 (6.9%) | 222 (0.6%) | <0.001 |
| Ischemic heart disease | 96 (24.7%) | 2,305 (6.0%) | <0.001 | 62 (15.9%) | 2,339 (6.1%) | <0.001 |
| Cerebrovascular disease | 26 (6.7%) | 866 (2.3%) | <0.001 | 23 (5.9%) | 869 (2.3%) | <0.001 |
| CMR parameters |  |  |  |  |  |  |
| LVEF (%) | 36.9 ± 5.1 | 59.8 ± 5.7 | <0.001 | 44.4 ± 9.5 | 59.7 ± 5.9 | <0.001 |
| RVEF (%) | 42.6 ± 10.2 | 57.4 ± 5.9 | <0.001 | 35.2 ± 5.7 | 57.5 ± 5.8 | <0.001 |
| AI-ECG scores |  |  |  |  |  |  |
| QCG-LVD | 0.28 ± 0.26 | 0.03 ± 0.06 | <0.001 | 0.16 ± 0.22 | 0.03 ± 0.06 | <0.001 |
| ECG-LVGLS | 14.44 ± 2.71 | 18.05 ± 1.05 | <0.001 | 15.60 ± 2.69 | 18.04 ± 1.09 | <0.001 |
| QCG-RVD | 0.14 ± 0.13 | 0.04 ± 0.05 | <0.001 | 0.14 ± 0.15 | 0.04 ± 0.05 | <0.001 |
| ECG-RVGLS | 18.56 ± 3.53 | 23.24 ± 1.49 | <0.001 | 19.18 ± 3.88 | 23.23 ± 1.50 | <0.001 |

LVEF : left ventricular ejection fraction
RVEF : right ventricular ejection fraction
QCG : quantitative electrocardiogram
LVD : left ventricle dysfunction
LVGLS : left ventricular global longitudinal strain
RVD : right ventricle dysfunction
RVGLS : right ventricular global longitudinal strain
LVH : left ventricular hypertrophy
LAE : left atrial enlargement

As expected, CMR-derived LVEF was significantly lower in the LVD group compared to those without LVD (36.9 ± 5.1% vs. 59.5 ± 6.1%; p < 0.001), and RVEF was markedly reduced in the RVD group (35.2 ± 5.7% vs. 57.4 ± 5.9%; p < 0.001). Corresponding AI-ECG scores were significantly higher among participants with ventricular dysfunction (all p < 0.001), highlighting the clear separation in AI-ECG values between normal and abnormal groups.

AI-ECG models demonstrated excellent performance in detecting LVD, with an area under the curve (AUC) of 0.887 (95% CI: 0.866–0.907) for Quantitative ECG (QCG™)-LVD and 0.896 (95% CI: 0.876–0.916) for ECG-LV global longitudinal strain (LVGLS). For RVD, AUCs were 0.778 (95% CI: 0.751–0.804) for QCG-RVD and 0.825 (95% CI: 0.800– 0.850) for ECG-RVGLS (Figure 2). Although performance for RVD detection was slightly lower than for LVD, the AI-ECG models still demonstrated substantial diagnostic value, particularly given the challenges of assessing RVD using ECG alone. In subgroup analyses, AI-ECG performance for LVD detection was particularly high in older individuals and males, suggesting potential demographic variations in model accuracy. Subgroup analyses revealed higher diagnostic accuracy for LVD in older individuals and males. For RVD, model performance remained consistent across subgroups stratified by body mass index (BMI) groups and individuals with and without comorbidities. Notably, the model exhibited slightly higher accuracy in subgroups with a higher prevalence of ventricular dysfunction, such as older individuals and those with ischemic heart disease. (Figure 3)

**Figure 1.**
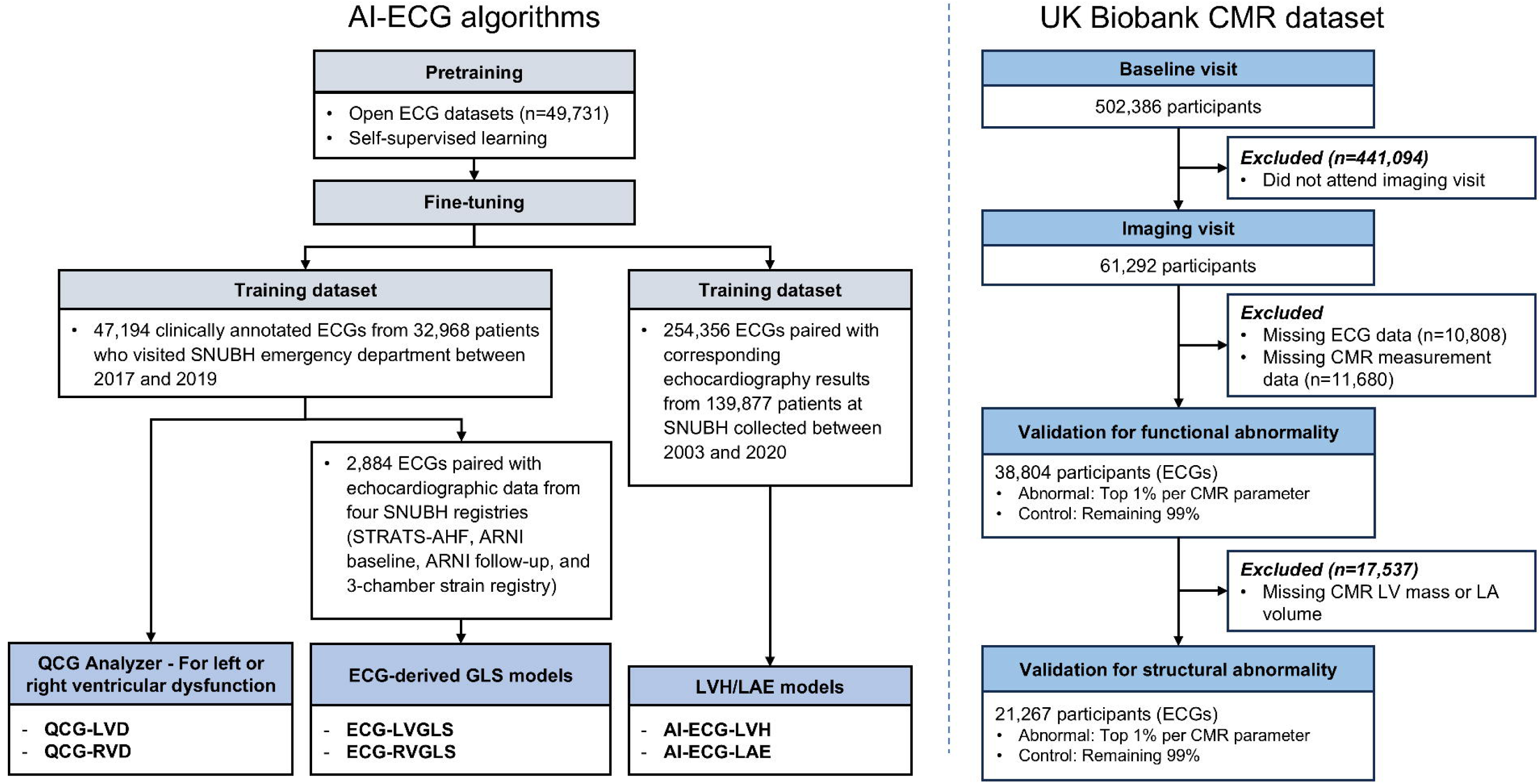
Flowchart illustrating AI-ECG algorithm development and validation dataset. The six AI-ECG models used in this study – four targeting functional abnormalities and two targeting structural abnormalities – were previously developed using ECG datasets from Seoul National University Bundang Hospital (SNUBH). These models were validated against CMR-derived values in the UK Biobank population. CMR = cardiac magnetic resonance imaging, ECG = electrocardiography, QCG = quantitative ECG, LVD = left ventricular dysfunction, RVD = right ventricular dysfunction, GLS = global longitudinal strain, LVH = left ventricular hypertrophy, LAE = left atrial enlargement.

**Figure 2.**
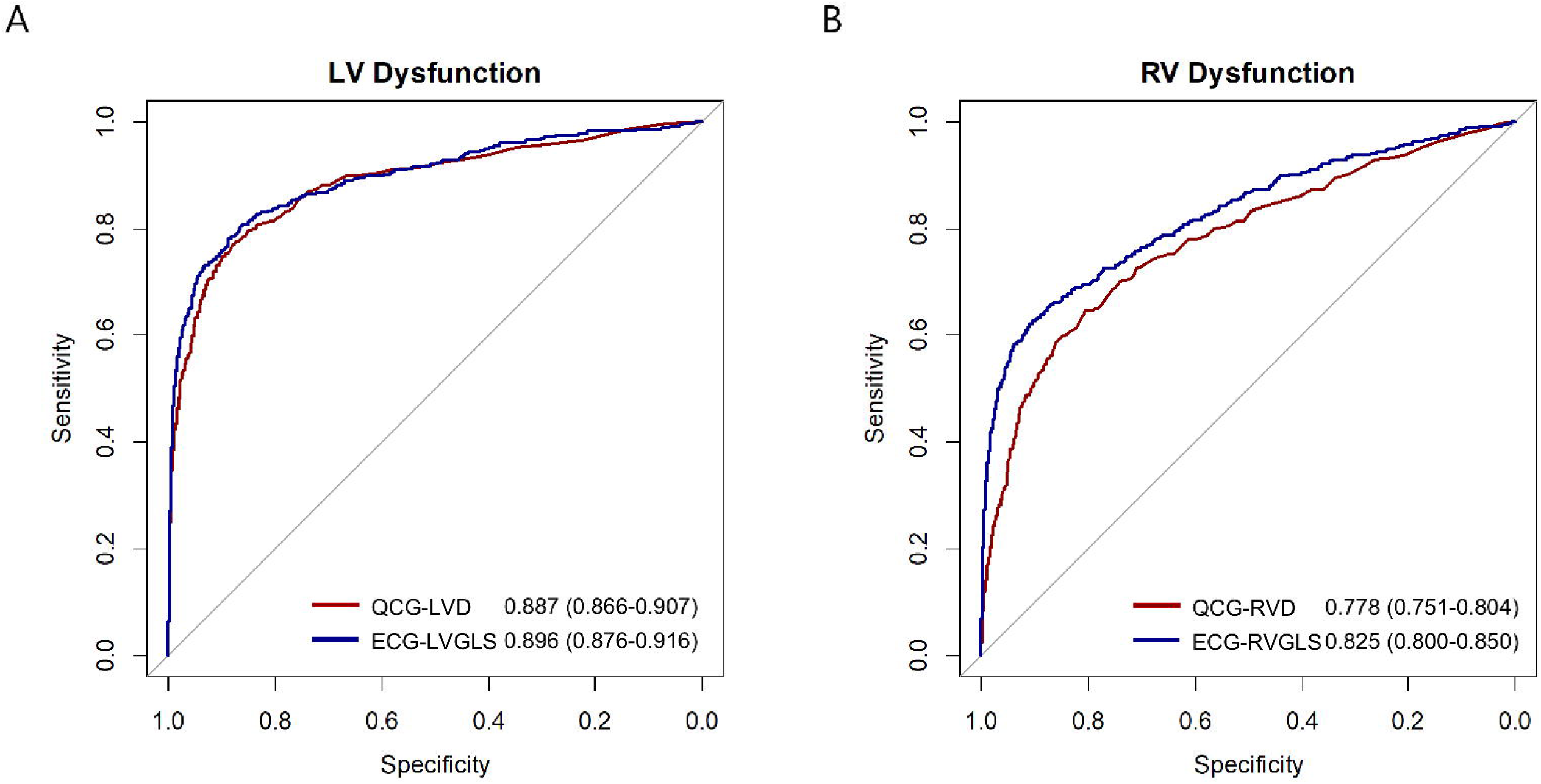
Diagnostic performance of AI-ECG for left and right ventricular dysfunction. Receiver operating characteristic (ROC) curves for AI-ECG models predicting (A) left and (B) right ventricular dysfunction, as defined by CMR-derived eft ventricular ejection fraction (LVEF) and right ventricular ejection fraction (RVEF), respectively.

**Figure 3.**
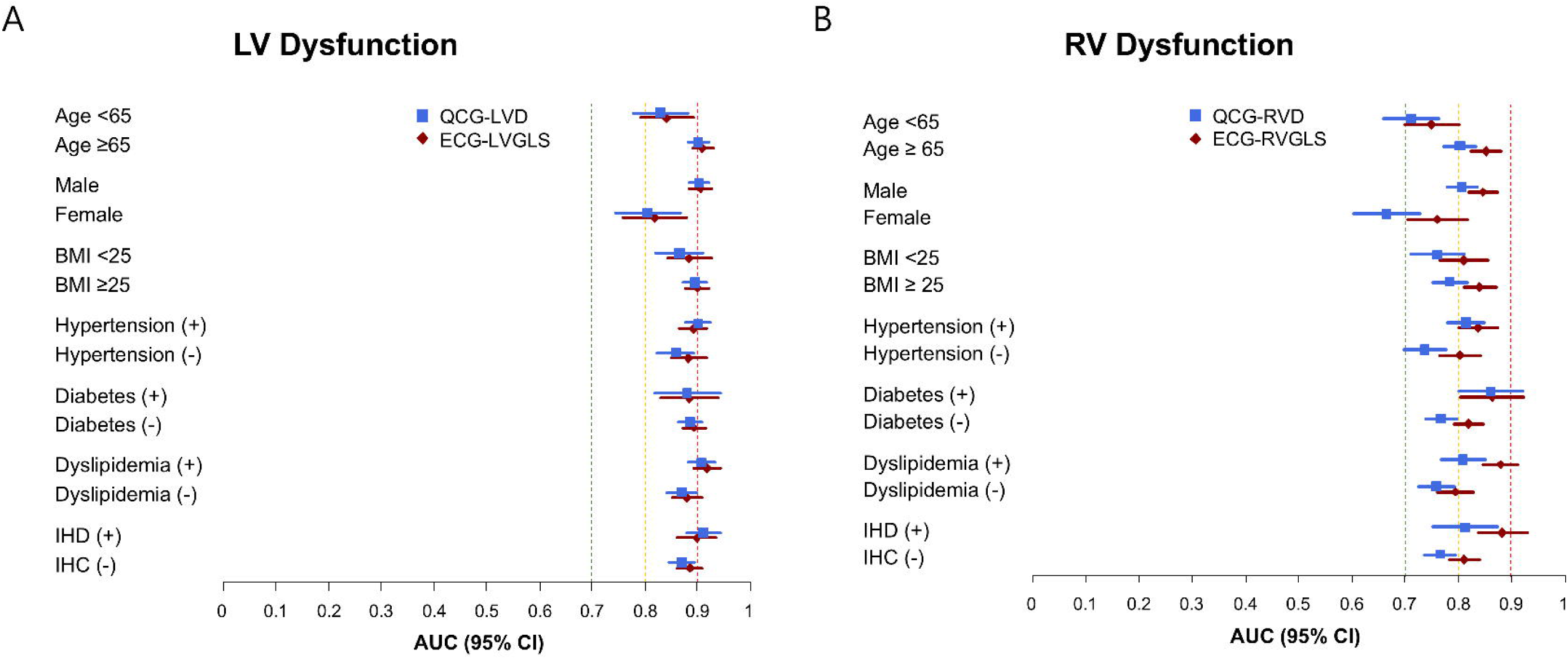
Subgroup analysis for ventricular dysfunction detection performances. (A) Forest plots showing AUC of QCG-LVD and ECG-LVGLS for LVD, and (B) QCG-RVD and ECG-RVGLS for RVD, across clinical subgroups stratified by age, sex, BMI, and comorbidities. AUC = area under the curve. Other abbreviations are as Figure 1.

Additionally, in the sub-cohort with available CMR-derived LVGLS data, the ECG-LVGLS model demonstrated robust performance in identifying abnormal LVGLS, defined as the lowest 1% of CMR-LVGLS distribution, with an AUC of 0.829 (95% CI, 0.794–0.865). The ECG-LVGLS was significantly lower in participants with abnormal CMR-LVGLS. (Supplementary Figure 1)

### AI-ECG Performance for Detecting Structural Abnormalities

Table 3 presents baseline characteristics stratified by the presence of left ventricular hypertrophy (LVH) and left atrial enlargement (LAE), as defined by CMR. Participants with LVH had a higher BMI (27.7 ± 5.3 vs. 26.4 ± 4.3 kg/m²; p < 0.001) and were more likely to have hypertension (55.4% vs. 31.0%; p < 0.001), heart failure (2.3% vs. 0.5%; p < 0.001), and ischemic heart disease (12.7% vs. 5.7%; p < 0.001). Similarly, individuals with LAE were older (67.6 ± 7.4 vs. 62.8 ± 7.5 years; p < 0.001), predominantly male (66.2% vs. 47.7%; p < 0.001), and had higher rates of hypertension (61.0% vs. 30.9%; p < 0.001) and ischemic heart disease (18.8% vs. 5.7%; p < 0.001).

**Table 3.**
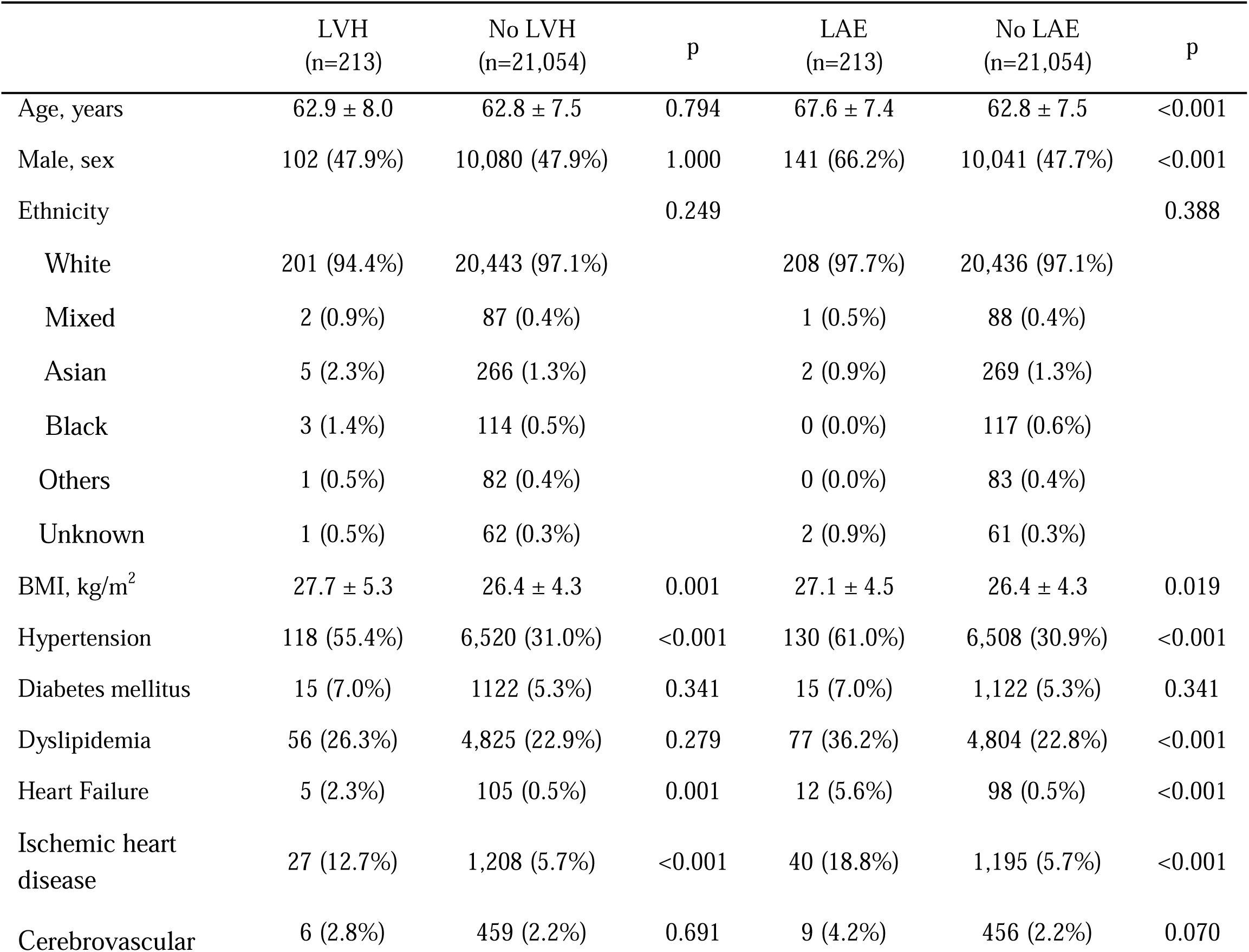
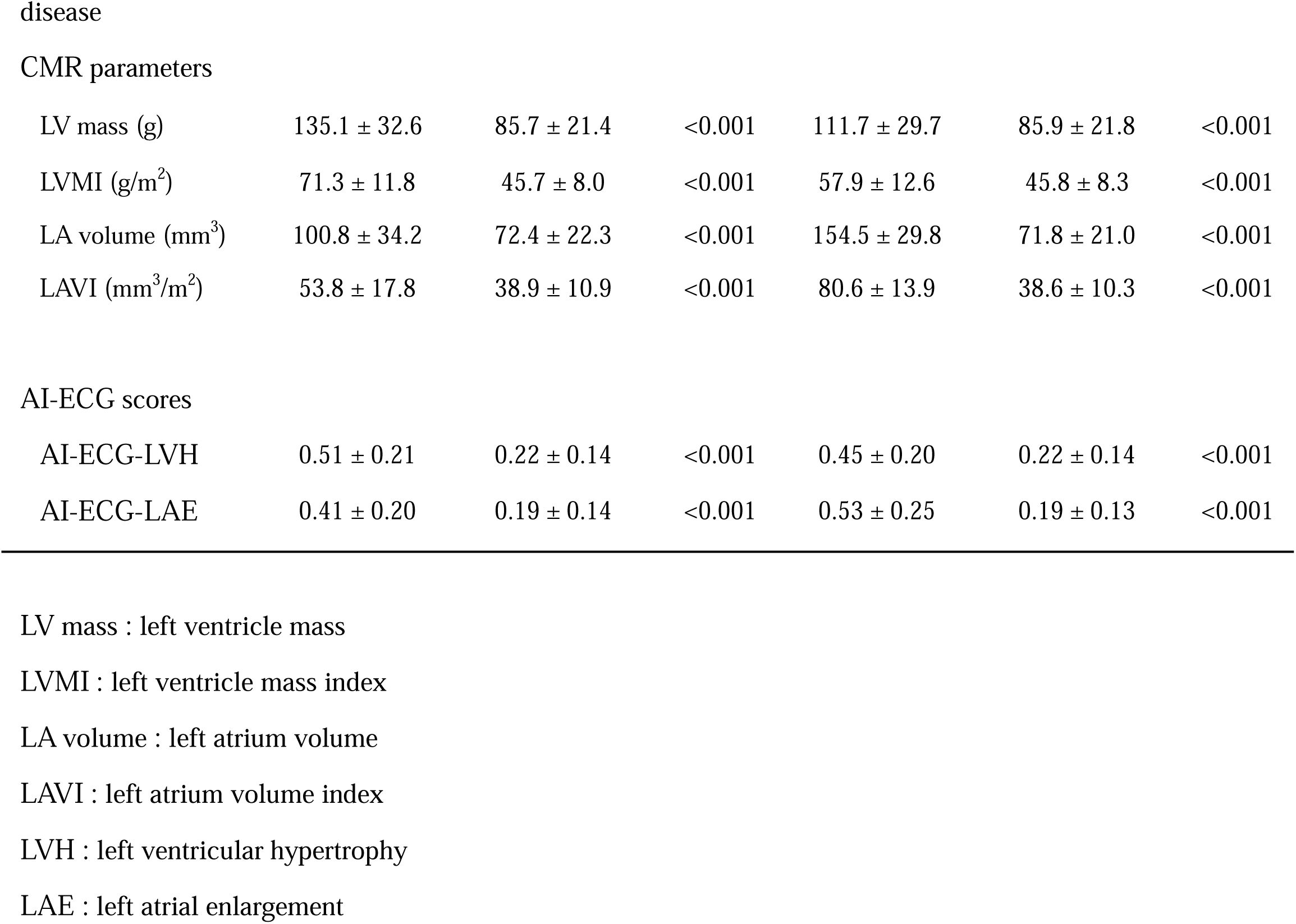
Baseline characteristics according to the presence of left ventricular hypertrophy and left atrial enlargement.

|  | LVH<br>(n=213) | No LVH<br>(n=21,054) | p | LAE<br>(n=213) | No LAE<br>(n=21,054) | p |
| --- | --- | --- | --- | --- | --- | --- |
| Age, years | 62.9 ± 8.0 | 62.8 ± 7.5 | 0.794 | 67.6 ± 7.4 | 62.8 ± 7.5 | <0.001 |
| Male, sex | 102 (47.9%) | 10,080 (47.9%) | 1.000 | 141 (66.2%) | 10,041 (47.7%) | <0.001 |
| Ethnicity |  |  | 0.249 |  |  | 0.388 |
| White | 201 (94.4%) | 20,443 (97.1%) |  | 208 (97.7%) | 20,436 (97.1%) |  |
| Mixed | 2 (0.9%) | 87 (0.4%) |  | 1 (0.5%) | 88 (0.4%) |  |
| Asian | 5 (2.3%) | 266 (1.3%) |  | 2 (0.9%) | 269 (1.3%) |  |
| Black | 3 (1.4%) | 114 (0.5%) |  | 0 (0.0%) | 117 (0.6%) |  |
| Others | 1 (0.5%) | 82 (0.4%) |  | 0 (0.0%) | 83 (0.4%) |  |
| Unknown | 1 (0.5%) | 62 (0.3%) |  | 2 (0.9%) | 61 (0.3%) |  |
| BMI, kg/m<sup>2</sup> | 27.7 ± 5.3 | 26.4 ± 4.3 | 0.001 | 27.1 ± 4.5 | 26.4 ± 4.3 | 0.019 |
| Hypertension | 118 (55.4%) | 6,520 (31.0%) | <0.001 | 130 (61.0%) | 6,508 (30.9%) | <0.001 |
| Diabetes mellitus | 15 (7.0%) | 1122 (5.3%) | 0.341 | 15 (7.0%) | 1,122 (5.3%) | 0.341 |
| Dyslipidemia | 56 (26.3%) | 4,825 (22.9%) | 0.279 | 77 (36.2%) | 4,804 (22.8%) | <0.001 |
| Heart Failure | 5 (2.3%) | 105 (0.5%) | 0.001 | 12 (5.6%) | 98 (0.5%) | <0.001 |
| Ischemic heart disease | 27 (12.7%) | 1,208 (5.7%) | <0.001 | 40 (18.8%) | 1,195 (5.7%) | <0.001 |
| Cerebrovascular | 6 (2.8%) | 459 (2.2%) | 0.691 | 9 (4.2%) | 456 (2.2%) | 0.070 |
| LV mass (g) | 135.1 ± 32.6 | 85.7 ± 21.4 | <0.001 | 111.7 ± 29.7 | 85.9 ± 21.8 | <0.001 |
| LVMI (g/m<sup>2</sup>) | 71.3 ± 11.8 | 45.7 ± 8.0 | <0.001 | 57.9 ± 12.6 | 45.8 ± 8.3 | <0.001 |
| LA volume (mm<sup>3</sup>) | 100.8 ± 34.2 | 72.4 ± 22.3 | <0.001 | 154.5 ± 29.8 | 71.8 ± 21.0 | <0.001 |
| LAVI (mm<sup>3</sup>/m<sup>2</sup>) | 53.8 ± 17.8 | 38.9 ± 10.9 | <0.001 | 80.6 ± 13.9 | 38.6 ± 10.3 | <0.001 |
| AI-ECG scores |  |  |  |  |  |  |
| AI-ECG-LVH | 0.51 ± 0.21 | 0.22 ± 0.14 | <0.001 | 0.45 ± 0.20 | 0.22 ± 0.14 | <0.001 |
| AI-ECG-LAE | 0.41 ± 0.20 | 0.19 ± 0.14 | <0.001 | 0.53 ± 0.25 | 0.19 ± 0.13 | <0.001 |

LV mass : left ventricle mass
LVMI : left ventricle mass index
LA volume : left atrium volume
LAVI : left atrium volume index
LVH : left ventricular hypertrophy
LAE : left atrial enlargement

**Table 4.** Prevalence of functional and structural abnormalities stratified by demographic and clinical characteristics.

|  | Total<br>(n=38,804) | LVD | RVD | Subgroup<br>(n=21,267) | LVH | LAE |
| --- | --- | --- | --- | --- | --- | --- |
| Age≥65 | 19,683 (50.7%) | 293 (1.5%) | 293 (1.5%) | 9,574 (45.0%) | 99 (1.0%) | 152 (1.6%) |
| Age<65 | 19,121 (49.3%) | 96 (0.5%) | 96 (0.5%) | 11,693 (55.0%) | 114 (1.0%) | 61 (0.5%) |
| Male | 18,709 (48.2%) | 308 (1.6%) | 295 (1.6%) | 10,182 (47.9%) | 102 (1.0%) | 141 (1.4%) |
| Female | 20,095 (51.8%) | 81 (0.4%) | 94 (0.5%) | 11,085 (52.1%) | 111 (1.0%) | 72 (0.6%) |
| BMI <25 | 15,348 (39.6%) | 118 (0.8%) | 118 (0.8%) | 8,510 (40.0%) | 75 (0.89%) | 73 (0.9%) |
| BMI ≥25 | 22,289 (60.4%) | 255 (1.1%) | 255 (1.1%) | 12,192 (60.0%) | 135 (1.1%) | 134 (1.1%) |
| Hypertension (+) | 12,778 (32.9%) | 213 (1.7%) | 194 (1.5%) | 6,638 (31.2%) | 118 (1.8%) | 130 (2.0%) |
| Hypertension (-) | 26,026 (67.1%) | 176 (0.7%) | 195 (0.7%) | 14,629 (68.8%) | 95 (0.7%) | 83 (0.6%) |
| Diabetes mellitus (+) | 2,275 (5.9%) | 50 (2.2%) | 44 (1.9%) | 1,137 (5.3%) | 15 (1.3%) | 15 (1.3%) |
| Diabetes mellitus (-) | 36,529 (94.1%) | 339 (0.9%) | 345 (0.9%) | 20,130 (94.7%) | 198 (1.0%) | 198 (1.0%) |
| Dyslipidemia (+) | 9,365 (24.1%) | 149 (1.6%) | 139 (1.5%) | 4,881 (23.0%) | 56 (1.2%) | 77 (1.6%) |
| Dyslipidemia (-) | 29,439 (75.9%) | 240 (0.8%) | 250 (0.8%) | 16,386 (77.0%) | 157 (1.0%) | 136 (0.8%) |
| Ischemic heart disease (+) | 2,401 (6.2%) | 96 (4.0%) | 62 (2.6%) | 1,235 (5.8%) | 27 (2.2%) | 40 (3.2%) |
| Ischemic heart disease (-) | 36,403 (93.8%) | 293 (0.8%) | 327 (0.9%) | 20,032 (94.2%) | 186 (0.9%) | 173 (0.9%) |

CMR-derived left ventricular mass index (LVMI) (76.6 ± 7.2 vs. 45.6 ± 7.9 g/m²; p < 0.001) and LAVI (80.6 ± 13.9 vs. 38.6 ± 10.3 mm³/m²; p < 0.001) were significantly elevated in LVH and LAE groups, respectively. AI-ECG scores were significantly elevated in participants with structural abnormalities. AI-ECG-LVH scores were markedly increased in participants with LVH (0.51 ± 0.21 vs. 0.22 ± 0.14; p<0.001), and AI-ECG-LAE scores were notably higher among those with LAE (0.53 ± 0.25 vs. 0.19 ± 0.13; p<0.001), demonstrating clear separation between groups.

AI-ECG demonstrated strong diagnostic accuracy for identifying structural abnormalities. AI-ECG-LVH achieved an AUC of 0.877 (95% CI, 0.851–0.902) for detecting LVH, while AI-ECG-LAE exhibited AUC of 0.883 (95% CI, 0.858–0.908) for detecting LAE (Figure 4). In subgroup analyses, predictive performance for LVH and LAE detection was robust across demographic categories, including age, sex, and BMI subgroups. The accuracy of AI-ECG was particularly strong in older individuals and males, reflecting trends similar to those observed in functional abnormality (ventricular dysfunction) analyses. Model performance was also enhanced in subgroups characterized by higher prevalence rates of cardiovascular comorbidities, notably hypertension and ischemic heart disease, highlighting the clinical relevance of AI-ECG as a potential non-invasive screening tool for structural cardiac abnormalities. (Figure 5)

**Figure 4.**
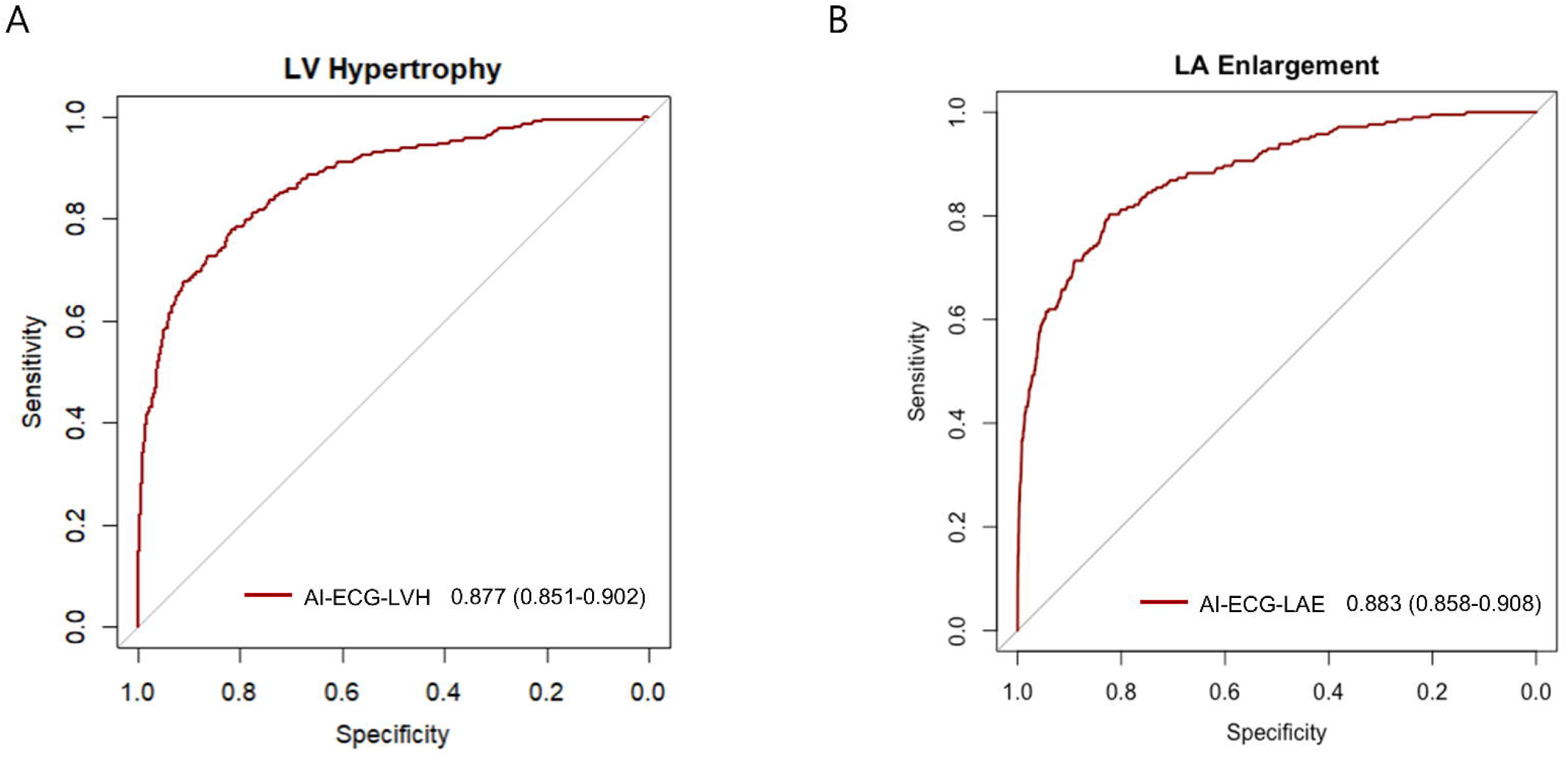
Diagnostic performance of AI-ECG for LVH and LAE. ROC curves demonstrating the predictive performance of AI-ECG models for detecting structural abnormalities: (A) LVH and (B) LAE defined by CMR. Abbreviations are as Figure 1 and 2.

**Figure 5.**
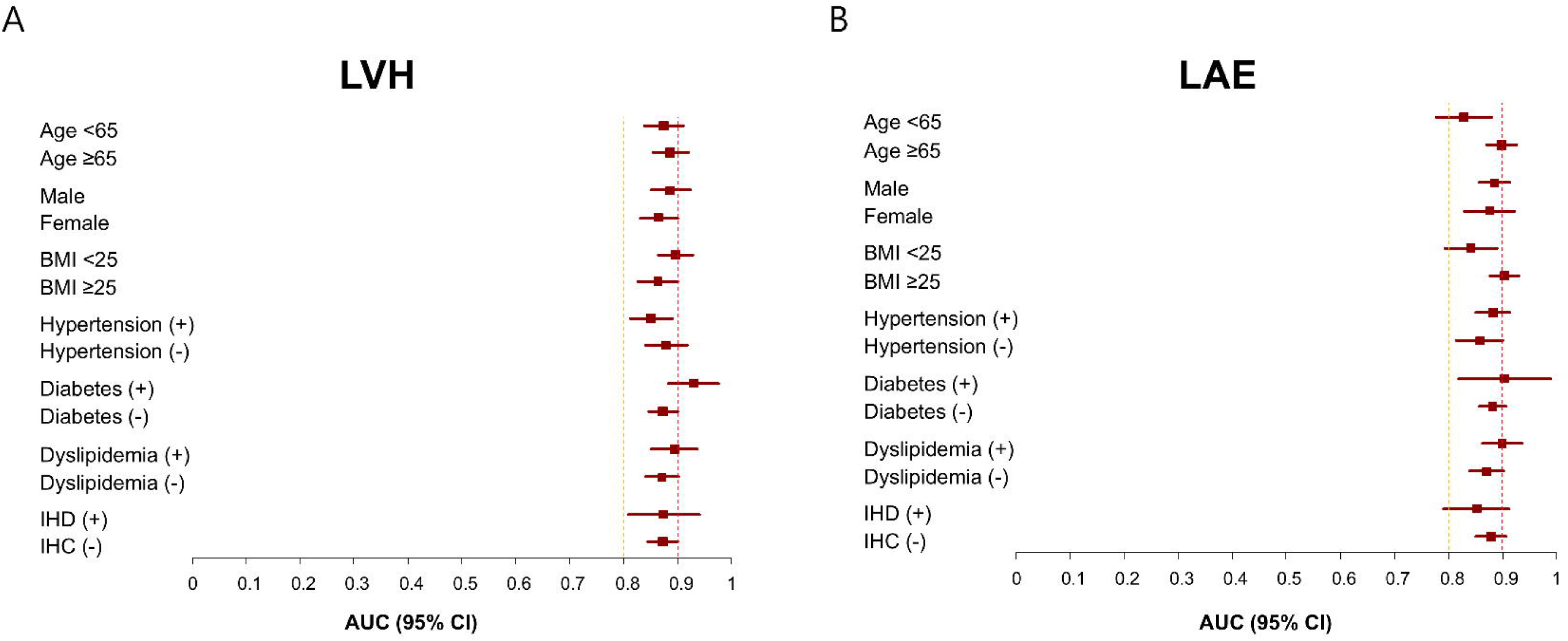
Subgroup analysis for structural abnormality detection. Forest plots showing (A) AUCs of AI-ECG-LVH and (B) AI-ECG-LAE across demographic and clinical subgroups. Abbreviations are as Figure 1 and 3.

## DISCUSSION

In this study of 38,804 UK Biobank participants, we demonstrated that AI-ECG can accurately detect structural and functional cardiac abnormalities, as defined by CMR. Using the top 1% of extreme CMR-derived values to define abnormalities, AI-ECG models achieved high diagnostic performance across a range of conditions: left ventricular dysfunction (AUC 0.887–0.896), right ventricular dysfunction (AUC 0.778–0.825), left ventricular hypertrophy (AUC 0.824), and left atrial enlargement (AUC 0.883). These findings support the potential utility of AI-ECG as a scalable, non-invasive tool for population-level cardiovascular screening, extending its application beyond the hospital-based Korean cohorts in which the models were originally developed.

Previous research has shown that AI-ECG can effectively identify various cardiac conditions. Notably, a Mayo Clinic study demonstrated reliable detection of left ventricular dysfunction using AI applied to ECGs, validated against echocardiography-defined ejection fraction.^8^ Our group has similarly developed ECG image–based models that perform well in detecting structural and functional abnormalities.^4,5^ However, prior studies have primarily focused on hospital-based or ethnically homogeneous populations. The present study uniquely validates these models in a multiethnic, community-based cohort from the UK Biobank, enhancing their generalizability and external validity. Despite differences in ethnicity and healthcare settings, our AI-ECG algorithms maintained high accuracy, suggesting the robust nature of ECG image-based analysis and generalizable convolutional neural network-based AI methodologies. This multiethnic validation significantly strengthens the external validity and potential clinical utility of AI-ECG for broad population-level screening.

CMR is widely regarded as the gold standard for evaluating cardiac structure and function due to its reproducibility and accuracy, particularly for parameters such as ventricular volumes and myocardial mass. Compared with echocardiography, CMR avoids geometric assumptions and operator dependency, resulting in more reliable assessments.^9^ Consequently, significant discrepancies between echocardiography and CMR-derived parameters have been consistently reported, with one study noting up to 60% discordance in patients with reduced LV ejection fraction (≤45%).^10–12^ This is especially important for right ventricular function, which is often difficult to evaluate via echocardiography due to its complex geometry and limited acoustic windows, making CMR a more reliable reference standard.^13^ Given these inherent methodological differences and the population-based nature of the UK Biobank cohort, we defined cardiac structural and functional abnormalities using the top 1% CMR-measured values rather than applying traditional echocardiographic thresholds. This approach enabled identification of individuals with marked deviations from normal cardiac function and provided a rigorous reference standard for AI-ECG validation in a general population setting.

Beyond confirming strong overall performance, our subgroup analyses offered additional insights into its performance across various demographic and clinical subgroups. Notably, diagnostic accuracy for both LVD and RVD was significantly higher among older participants and males compared to younger individuals and females. The presence of hypertension significantly enhanced the diagnostic accuracy of AI-ECG for both LVD and RVD, while diabetes and dyslipidemia tended to improve RVD detection. Regarding structural abnormalities, the diagnostic accuracy of AI-ECG for LVH was greater in participants with diabetes, whereas the predictive performance for LAE was enhanced among older individuals and those with higher BMI. These trends likely reflect the greater degree of cardiac remodeling in individuals with established cardiovascular risk factors, resulting in more pronounced ECG changes and improved model discrimination. Although performance was broadly consistent across subgroups, these findings suggest that AI-ECG may have particularly high clinical value in high-risk populations.

The robust diagnostic performance and high accessibility of AI-ECG demonstrated in this study suggest substantial implications for clinical practice. In real-world healthcare settings, AI-ECG could serve as a scalable, non-invasive initial screening tool, efficiently identifying individuals at higher cardiovascular risk who may benefit from more detailed diagnostic assessments or preventive interventions. For instance, AI-ECG could be integrated into routine primary care visits or community-based screening programs, promptly flagging patients with subtle but clinically meaningful abnormalities such as early-stage heart failure, asymptomatic left ventricular hypertrophy, or atrial remodeling indicative of increased atrial fibrillation risk. This early identification could facilitate timely intervention, potentially reducing morbidity and mortality associated with undetected cardiac conditions. Furthermore, because our AI-ECG algorithms operate on standard printed ECG images, their implementation requires minimal infrastructural changes, allowing rapid integration into diverse healthcare environments, including resource-limited settings where access to advanced cardiac imaging modalities remains challenging. Ultimately, our results highlight the practical and clinical value of incorporating AI-ECG into broader cardiovascular disease prevention and management strategies, emphasizing its potential to improve patient outcomes on a population scale.

This study has several limitations. While the UK Biobank dataset expands research from Korean populations to a predominantly European cohort, its primarily White demographic composition may still limit the generalizability, highlighting the need for validation in more diverse multi-ethnic cohorts. The top 1% CMR values offers reasonable and practical cut-off value for defining the functional and structural abnormalities in general population, but may not align with previously established thresholds, potentially limiting its application in routine clinical workflows where fixed cut-offs are standard. The black-box nature of AI-ECG models may pose challenges for clinical adoption, underscoring the need for further efforts to improve interpretability. Lastly, while our AI-ECG algorithms demonstrated robust diagnostic performance, whether such algorithms can be effectively integrated into real-world clinical practice to improve patient care remains to be determined. Future prospective studies are needed to evaluate the clinical utility of AI-ECG as a screening tool, and to confirm whether its application can meaningfully enhance clinical decision-making and improve cardiovascular outcomes.

In conclusion, the newly developed AI-ECG scores are highly effective in detecting functional and structural cardiac abnormalities within a general population. These findings highlight AI-ECG’s potential as a cost-effective and accessible screening tool, addressing limitations in availability and diagnostic accuracy associated with current imaging modalities, such as echocardiography and CMR.

## Methods

### Study population

This study utilized data from the UK Biobank, a large-scale, prospective cohort comprising over 500,000 participants aged 40 to 69 years, recruited between 2006 and 2010. The cohort includes data collected from hospital records and health check-ups in the general population. Detailed protocols for recruitment and imaging have been previously described.^14^ Among the 502,386 participants, 61,292 underwent imaging visits that included both ECG and CMR. We excluded 10,808 participants without ECG data and 11,680 without available CMR parameters, yielding a final study population of 38,804 individuals (Figure 1). A sub-cohort of 21,267 participants with complete structural CMR measurements, specifically left ventricular mass and left atrial volume, was used for the evaluation of structural abnormalities. This sub-cohort also included CMR-derived left ventricular global longitudinal strain (LVGLS) data, allowing additional validation of AI-ECG models targeting myocardial strain. This study was conducted under the ethical approval granted to the UK Biobank by the National Health Service National Research Ethics Service (Ref 11/NW/0382, extended under Ref 16/NW/0274). Access to anonymized participant data was granted through an approved UK Biobank application.

### AI algorithms

We evaluated six previously developed AI-ECG scores: four targeting functional abnormalities (QCG-LVD for left ventricular dysfunction, QCG-RVD for right ventricular dysfunction, ECG-LVGLS for LV global longitudinal strain, and ECG-RVGLS for RV global longitudinal strain) and two targeting structural abnormalities (AI-ECG-LVH for LV hypertrophy and AI-ECG-LAE for LA enlargement). These models are based on modified convolutional neural networks (CNNs) that utilize a shared encoder and task-specific output layers to analyze ECG signals or images.

The QCG-LVD and QCG-RVD models were developed as part of the QCG™ framework and are embedded in the ‘ECG Buddy’ software, which received regulatory approval from the Korean Ministry of Food and Drug Safety (MFDS) in January 2024. The encoder component was pretrained on 49,731 public ECG datasets via self-supervised learning and fine-tuned using 47,194 annotated ECG images from 32,968 patients at Seoul National University Bundang Hospital between 2017 and 2019. This multi-task learning framework enabled the models to classify rhythms and predict critical cardiovascular conditions such as shock, cardiac arrest, acute coronary syndrome, and heart failure.^4,15,16^ The ECG-LVGLS and ECG-RVGLS models were developed using a transfer learning approach. The CNN encoder from the QCG system was retained, while new task-specific networks were trained to output ECG-based global longitudinal strain estimates. Training utilized data from 2,882 patients across four Korean hospital cohorts, with external validation in an independent sample.^5^ The AI-ECG-LVH and AI-ECG-LAE models share a similar CNN architecture and were trained on 254,356 standard 12-lead ECGs matched with echocardiography results from 139,877 patients at Seoul National University Bundang Hospital between 2003 and 2020. This study adhered to the principles of the Declaration of Helsinki and was approved by the institutional review board (IRB No. B-2306-832-101).

### AI analysis of ECG

Resting ECGs (RestECG, Field 20205) were obtained from participants during initial imaging (instance 2), or repeat imaging (instance 3) visits and were provided by the UK Biobank in XML format (https://biobank.ndph.ox.ac.uk/showcase/field.cgi?id=20205).

Voltage signals for the 12 standard leads (I, II, III, aVR, aVL, aVF, V1–V6) were extracted by parsing each <WAVEFORMDATA> element. The <Resolution units=“uVperLsb”>5</RESOLUTION> tag indicates that each integer tick represents 5 µV. Therefore, the raw values were multiplied by 5 to convert them to µV and then divided by 1,000 to express them in mV. The resulting 10-second recordings sampled at 500 Hz (5,000 samples per lead) were split into four consecutive 2.5-second segments and arranged in the first three rows of a 4×4 grid. The fourth row displays the full 10-second Lead II tracing to replicate a standard 12-lead ECG printout layout. These processed signals were then transformed as standard 12-lead ECG report images and served as input to the AI models to generate the six AI-ECG scores evaluated in this study.

### CMR image acquisition and parameter extraction

CMR imaging in the UK Biobank was conducted using 1.5-Tesla MAGNETOM Aera scanners (Siemens Healthcare, Syngo Platform VD13A), following a previously established protocol.^17^ Quantitative cardiac and aortic parameters were extracted using a validated automated analysis pipeline powered by machine learning.^18^ This pipeline, based on convolutional neural networks, performed segmentation of cardiac and aortic structures from short-axis, long-axis, and cine images. Specifically, short-axis cine images were used to segment the ventricles and myocardium through a fully convolutional neural network trained on expert-labeled data from 3,975 individuals. This enabled the computation of ventricular volumes, myocardial mass, and ejection fractions. Comprehensive morpho-functional measurements were derived for all four cardiac chambers and two aortic segments. To account for body size, all volume-based measurements were indexed to body surface area.

### Definitions

In this study, we defined abnormalities in CMR parameters to assess various aspects of cardiac function and structures, indicative of underlying cardiovascular conditions. We utilized preprocessed CMR measurements provided by the UK Biobank imaging project, which were generated through standardized imaging protocols and automated post-processing pipelines developed and validated in prior studies.^17,18^

Specifically, we focused on LVD, RVD, LVH, and LAE. LVD and RVD were defined using CMR-derived LVEF, RVEF, or global longitudinal strain (LVGLS and RVGLS), respectively. LVH and LAE were defined based on left ventricular mass and left atrial maximum volume, respectively, both indexed to body surface area.

To account for population-based variability, we defined abnormalities as the top 1% of values for each CMR-derived parameter. For LVH, the cutoff was determined separately for males and females, reflecting sex-specific differences in normative values of LV mass index, as commonly applied in echocardiographic criteria.^19^ The specific cutoff values corresponding to the top 1% for each parameter are detailed in Supplementary Table 1. The performance of the AI-ECG algorithm in detecting these defined abnormalities were evaluated, aiming to determine its effectiveness in identifying early markers of cardiovascular dysfunction and its potential applicability in a clinical context.

### Statistical analysis

The predictive performance of AI-ECG models was assessed using sensitivity, specificity, and the area under the receiver operating characteristic curve (AUROC). Continuous variables were reported as mean ± standard deviation, and categorical variables as counts and percentages. Logistic regression models were used to adjust for potential confounders. A p-value < 0.05 was considered statistically significant. All analyses were performed using R (version 4.4.1; https://www.R-project.org).

### Conflicts of interest

Joonghee Kim, MD, PhD developed the algorithm. He also founded a start-up company, ARPI Inc., where he serves as the CEO. Youngjin Cho, MD, PhD works for the company as a research director. Haemin Lee works for the company as data scientist. Otherwise, there is no conflict of interest for the other authors

## Data Availability

https://www.ukbiobank.ac.uk/

## Acknowledgement

This research was supported by a grant of the Korea Health Technology R&D Project through the Korea Health Industry Development Institute (KHIDI), funded by the Ministry of Health & Welfare, Republic of Korea (grant number: RS-2023-00265933)

---

**Supple Fig 1.**
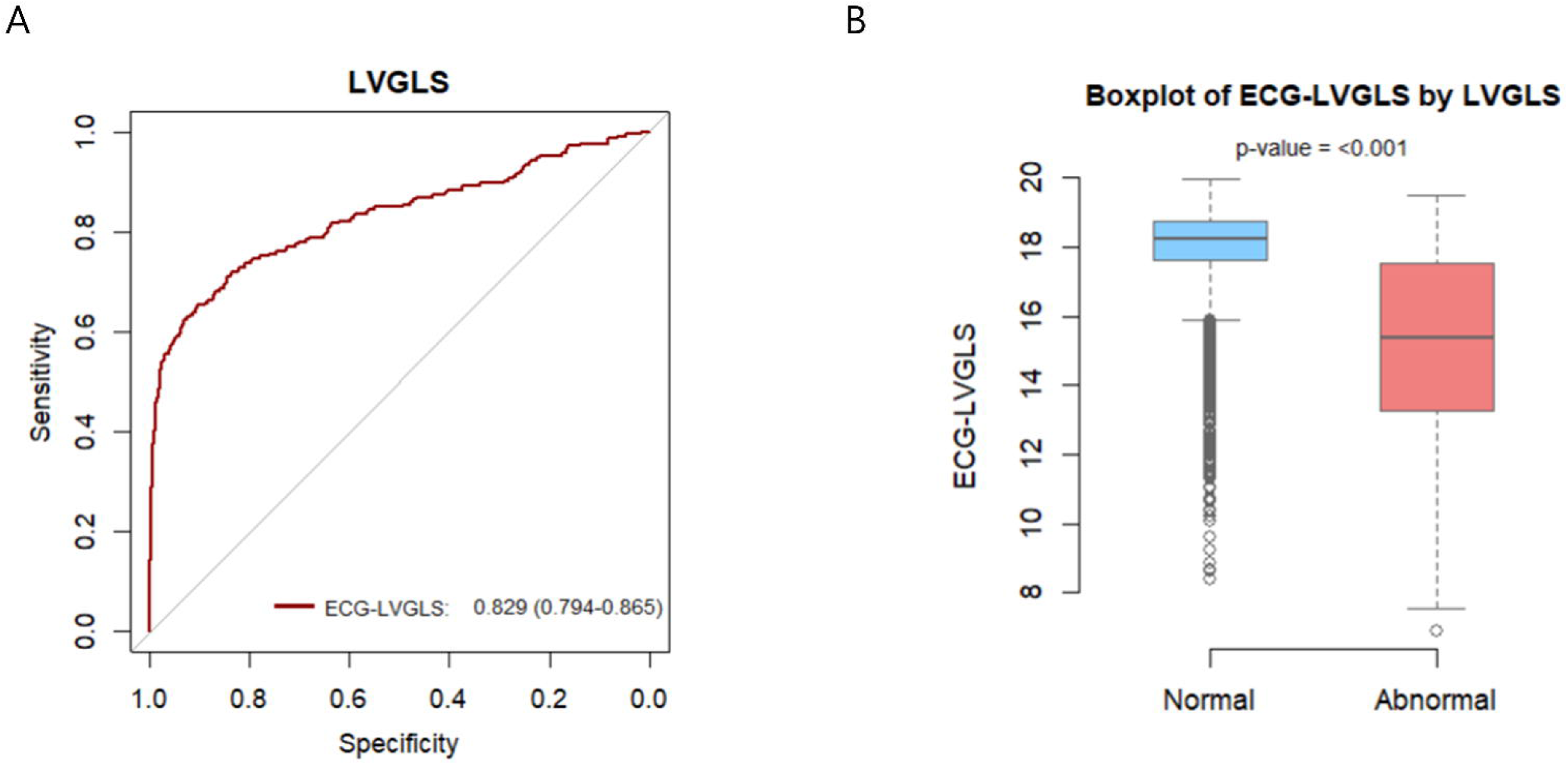

## Notes

### Author Declarations

This study adhered to the principles of the Declaration of Helsinki and was approved by the institutional review board (IRB No. B-2306-832-101).

